# A COVID-19 Infection Risk Model for Frontline Health Care Workers

**DOI:** 10.1101/2020.03.27.20045336

**Authors:** Louie Florendo Dy, Jomar Fajardo Rabajante

## Abstract

The number of confirmed COVID-19 cases admitted in hospitals is continuously increasing in the Philippines. Frontline health care workers are faced with imminent risks of getting infected. In this study, we formulate a theoretical model to calculate the risk of being infected in health care facilities considering the following factors: the average number of encounters with a suspected COVID-19 patient per hour; interaction time for each encounter; work shift duration or exposure time; crowd density, which may depend on the amount of space available in a given location; and availability and effectiveness of protective gears and facilities provided for the frontline health care workers. Based on the simulation results, a set of risk assessment criteria is proposed to classify risks as ‘low’, ‘moderate’, or ‘high’. We recommend the following: (i) decrease the rate of patient encounter per frontline health care worker, e.g., maximum of three encounters per hour in a 12-hour work shift duration; (ii) decrease the interaction time between the frontline health care worker and the patients, e.g., less than 40 minutes for the whole day; (iii) increase the clean and safe space for social distancing, e.g., maximum of 10% crowd density, and if possible, implement compartmentalization of patients; and/or (iv) provide effective protective gears and facilities, e.g., 95% effective, that the frontline health care workers can use during their shift. Moreover, the formulated model can be used for other similar scenarios, such as identifying infection risk in public transportation, school classroom settings, offices, and mass gatherings.

## Introduction

As of March 20, 2020, Coronavirus Disease (COVID-19) has infected 250,704 worldwide, resulting in 10,256 deaths, with Italy surpassing China in the reported number of deaths on the same day [1,2]. Aggressive suppression strategies have been recommended [3], and countries across the world have implemented strategies to mitigate the damage caused by this pandemic [4].

Health care workers work in the frontlines across the world unceasingly, running the greatest risk of getting infected and infecting others in their immediate environment – in the hospital, at home – or wherever they go. In the Philippine context, health care system can be described as a two-tiered [5]. There is a huge disparity in the capacity between the public and private health sectors [5]. As of 2016, there are a total of 101,688 hospital beds, with a ratio of 23 beds for 10,000 people in the National Capital Region, and more than half (53.4%) of these are in private hospitals [5]. The number of confirmed COVID-19 cases admitted in hospitals is continuously increasing exponentially [6]. Therefore, given that the health system is likely to be overwhelmed [5,7], these frontline health care workers (frontliners) are faced with unimaginable risks of getting infected.

Every doctor, nurse, medical technologist, radiation technologist, nursing assistant, hospital janitor and security guard will inevitably face the risk of COVID-19 infection. Here, we formulate a mathematical model to investigate how many frontliners are expected to be infected under certain scenarios [8]. We use this expected number of possible new infections as a measure of the risk. A set of risk assessment criteria has been formulated based on the theoretical results to determine if a frontliner has low, moderate or high risk of COVID-19 infection.

Mathematical models can be used for predicting scenarios and in prescribing solutions to problems [8,9,10,11], such as addressing the COVID-19 pandemic. Based on the simulations and risk assessment, several recommendations are suggested to inhibit the spread of SARS-CoV-2, especially in health care facilities.

## Results and Discussion

The formulated risk model (see Appendix: Methods) aims to examine the risk factors of virus transmission per day, quantify these risks, estimate the number of new infections, and suggest ways to minimize these risks. There are several factors that determine the risk of infection:

- Average number of COVID-19 patients (or, in other settings, number of susceptible persons) entering a given location at a given time, whether they are confirmed to be positive or not;
- Average number of encounters with a patient (or, in other settings, any susceptible person) at a given time, whether COVID-19 infected or not, where an encounter is defined to be less than or equal to 30 seconds;
- Duration of interaction of each of these encounters;
- Work shift duration of each frontliner (or, in other settings, can be equivalent to the exposure time for any person in public transportation, offices, classrooms, and mass gatherings);
- Crowd density, which may depend on the amount of space available in a given location, the presence of compartments or dividers in a room, and how frequent cleaning is done in the environment as the density of SARS-CoV-2 virus particles present on surfaces limits the safe space available; and
- Level of protection present (e.g., isolation booths, N95 masks, face and eye shields) including level of protection to reduce exposure to aerosolized particles (e.g., for those tasked to do intubation either via direct or via video laryngoscopy, to do nasopharyngeal or oropharyngeal swabs).

In the following discussion, the parameters will be discussed in terms of the frontliner setting, but the parameters can likewise be applied in other settings, such as in crowded places, classrooms, offices, public vehicles, and markets.

### Average Number of Encounters and Work Shift Duration (Exposure Time)

The average number of encounters per hour can be defined as the average number of patients a frontliner has interacted in an hour given that an interaction is less than or equal to 30 seconds. We can convert number of patients per minute to number of encounters using the following formula:

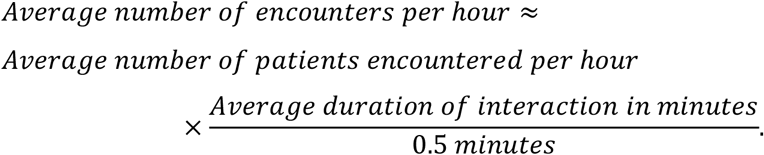

For example, a 10-15 minute interview with a patient is equivalent to 20-30 encounters. A doctor doing brief rounds on 10 patients (where each patient is looked upon in at most 30 seconds) or the situation of having 5 patients per minute are equivalent to 10 encounters.

According to our simulation results, the expected number of infected frontliners increases as the average number of encounters between the frontliner and COVID-19 patients increases as well as when work shift duration or exposure time increases (Figure 1A & 1B). If there is highly interacting population (e.g., the average number of encounters per hour is 120, which means that the frontliners and the patients are interacting every 30 seconds) or a series of long interactions (e.g., 4 patient interviews per hour where each interview takes 15 minutes), then there is a high chance that one person will be infected. If there is low interaction rate (e.g., seeing one patient only for less than or equal to 30 seconds once per hour), then the chance of getting infected is low but the risk is not zero.

**Figure 1.**
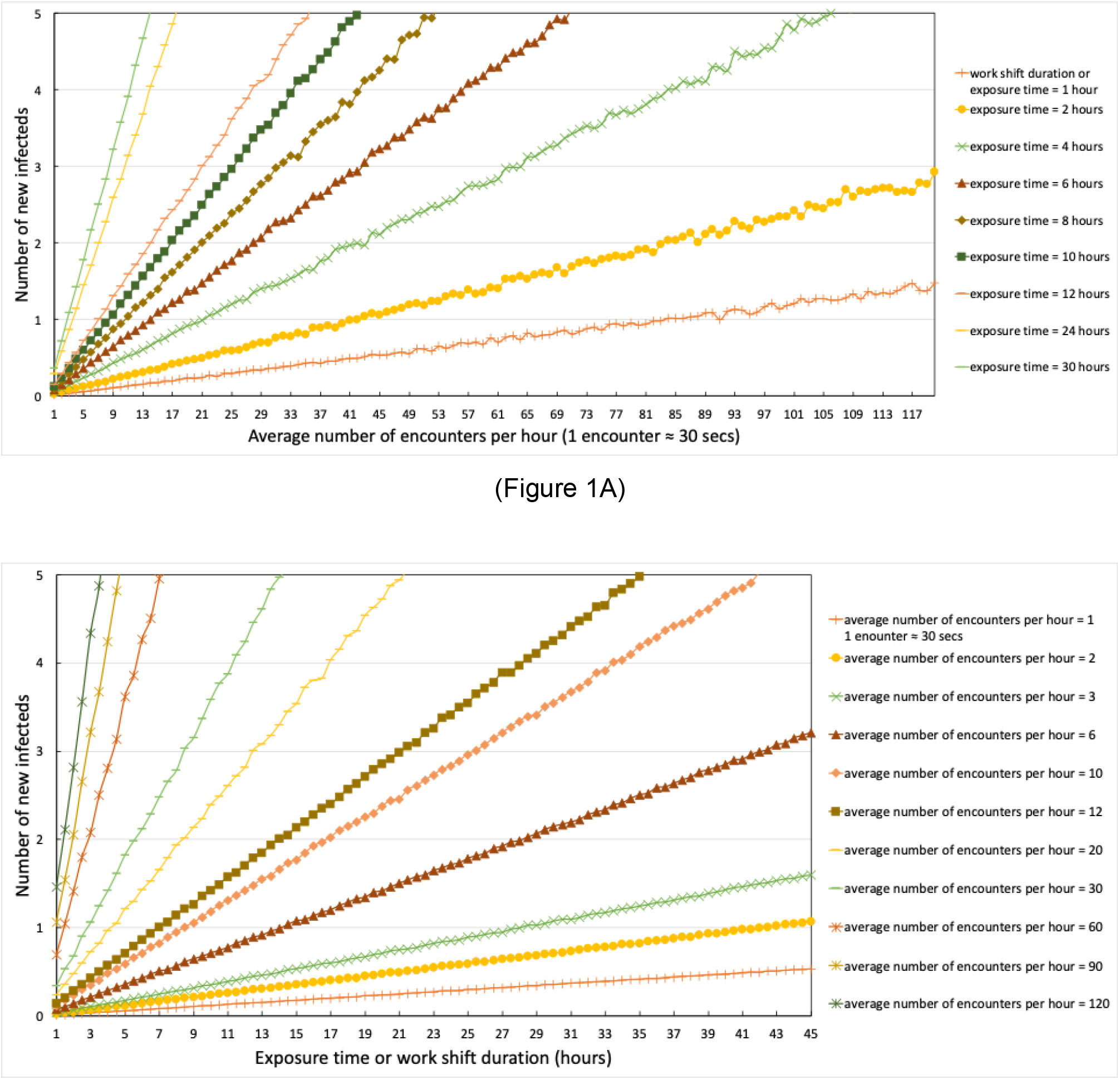
Relationship of work shift duration or exposure time, average number of encounters per hour, and the relative risk of a frontliner getting infected, which is proportional to the potential number of newly infected. Parameters used: *S*_0_=100, *S*_*max*_=100, no protection, *I*_0_=1. (**A**) Relationship between average number of encounters per hour and expected number of new infecteds. (**B**) Relationship between work shift duration or exposure time and expected number of new infecteds.

It should be noted that if the number of possible infected frontliners is greater than or equal to one, then there is high risk of infection; if the number of possible infected frontliners is less than one but not equal to zero, then there is still some level of risk (low or moderate risk of infection). In a 7-hour work shift duration, there is high chance a frontliner will be infected if the interaction rate is around 12 encounters per hour (Figure 1B). This can be imagined as a triage nurse seeing 1 patient for at most 30 seconds every 5 minutes during the duration of his or her 7-hour work shift.

The number of newly infected frontliners is directly proportional to the average number of encounters per hour (Table 1). Regardless of work shift duration, a hospital security guard or a triage nurse entertaining 120 persons per hour (1 patient every 30 seconds) is at least 12 times more likely to get infected than a medical or radiation technologist encountering 10 patients per hour. A doctor conducting long duration interviews and examinations on patients (20-80 persons per hour) is 2 to 8 times more likely to get infected than for example, a radiation technologist.

**Table 1.** Relative risk compared to 10 encounters per hour.

| Number of encounters per hour | Relative risk compared to 10 encounters per hour | Assigned Risk Points* | Examples |
| --- | --- | --- | --- |
| 120 | 12x | 10 | Security guard, triage nurse, doctor doing long interviews |
| 100 | 10x | 9.5 |  |
| 80 | 8x | 9 | A doctor conducting moderate to long duration interviews and examinations on several patients |
| 60 | 6x | 8 |  |
| 40 | 4x | 7 |  |
| 30 | 3x | 6.5 |  |
| 20 | 2x | 6 |  |
| 10 | 1x | 5 | Radiologic technologist who does chest X-rays for persons-under-investigation (PUIs), or anyone who takes nasopharyngeal or oropharyngeal swab |
| 6 | 0.5x | 4 |  |
| 3 | 0.25x | 3 |  |
| 2 | 0.167x | 2 |  |
| 1 | 0.0833x | 1 | Medical technologist who collects specimen only at certain timeslots, or any health worker with minimal patient interaction |

\*this is a sample qualitative point system that can be adjusted or modified depending on the situation.

Regardless of the type of frontliner, a work shift duration of at least 10 hours is at least 1.25 times more likely to be infected than that of 8 hours. The longer the work shift duration or exposure time, the higher the infection risk (Table 2).

**Table 2.** Relative risk compared to an 8-hour work shift duration or exposure time.

| Work shift duration or exposure time | Relative risk compared to an 8-hour work shift duration or exposure time | Assigned Risk Points* |
| --- | --- | --- |
| 30 | 3.75x | 10 |
| 24 | 3x | 8 |
| 12 | 1.5x | 6 |
| 10 | 1.25x | 3 |
| 8 | 1x | 2.5 |
| 6 | 0.75x | 2 |
| 4 | 0.5x | 1.5 |
| 2 | 0.25x | 1 |
| 1 | 0.125x | 0.5 |

\*this is a sample qualitative point system that can be adjusted or modified depending on the situation.

### Crowd Density

We define crowd density as the number of people in a room divided by the maximum capacity of the room. We can also define it as the average proportion of COVID-19 infected entities present within a 2-meter radius (minimum radius for social distancing) from the health care worker. The entity may be an infected patient (confirmed or not confirmed), or any object or surface in the immediate environment that contains SARS-CoV-2 virus particles. There is evidence that the virus particles stay as long as 3 hours as aerosols and 72 hours on plastic surfaces [12].

Looking at the following figure (Figure 2), crowd density acts as a fraction that modifies the risk of getting infected at all levels of encounter rates, from the laboratory (low encounter rate per hour) to the triage area (high encounter rate per hour). A crowded place where social distancing is not highly implemented can initiate transmission of the disease. Having more space available for each patient, putting dividers between infected patients, and cleaning the workspaces more often lead to a lower crowd density. A lower crowd density implies that the frontliner is receiving a lesser fraction of the risk of infection. A higher crowd density increases the chance of being infected.

**Figure 2.**
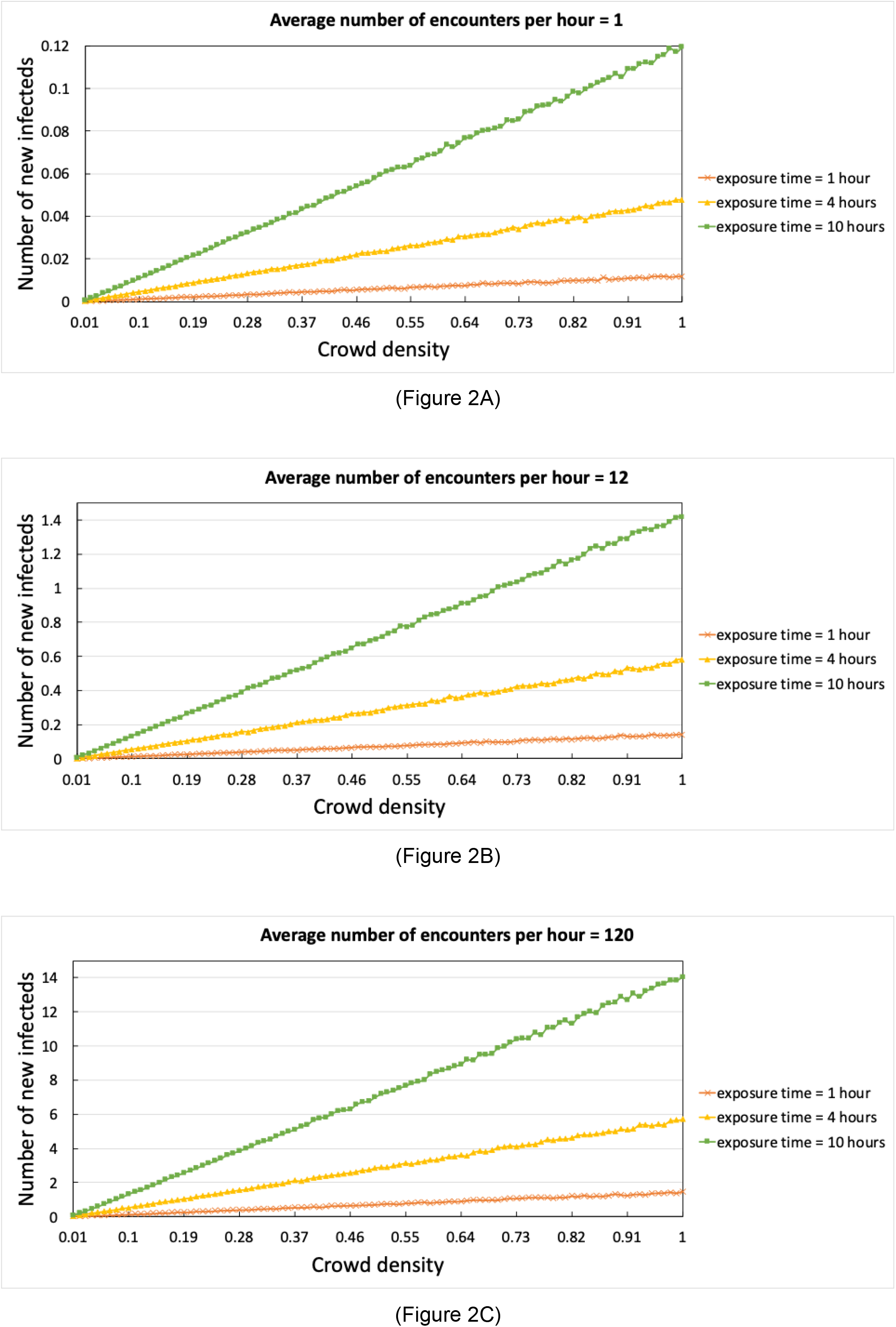
Effect of crowd density on infection risk with varying number of encounters and exposure time (work shift duration). Parameters used: *S*_0_=100, *S*_*max*_=100, no protection, *I*_0_=1. Average number of encounters per hour (**A**) = 1, (**B**) = 12, (**C**) = 120.

### Initial Number of Infected Patients

The effect of crowd density is so important that, even if ten COVID-19 infected patients enter the same room at the same time, the risk of the frontliner getting infected can be dramatically reduced by reducing the crowd density (Figure 3). For example, a health care worker in a room with crowd density of 10% is at least 95% less likely to be infected than a health care worker in a room with crowd density of 100%. Moreover, as expected, the number of newly infected patients in a room is directly proportional to the infection risk faced by the frontliners (Figure 3).

**Figure 3.**
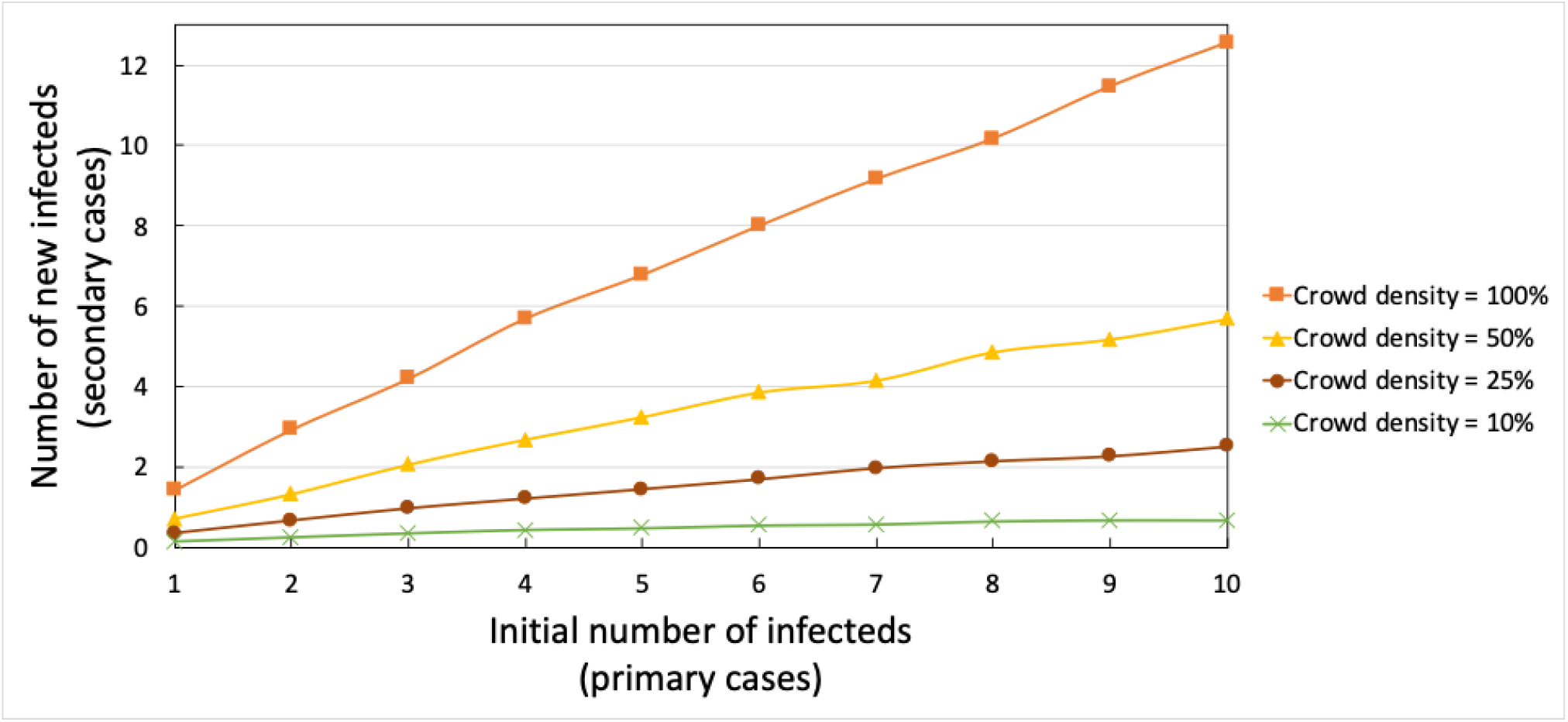
Risk of infection determined by the number of infected cases present in a room with the frontliners. Parameters used: *S*_*max*_=100, no protection, average number of encounters per hour = 120, work shift duration or exposure time = 1 hour.

### Protection Level

Protection level is defined as the fraction of the risk being removed or mitigated by measures done by the health care worker or any other person. It has a minimum value of 0 and a maximum value of 1. It can be observed in Figure 4 that a 95% or better protection level significantly reduces infection risk. We can assign values which may be additive as shown in Table 3.

**Table 3.** Examples of Protection Level Points for every procedure and equipment worn.

| Protection Level<br>Points* | Description |
| --- | --- |
| 0.00 | Having no personal protective equipment (PPE), not following hand hygiene |
| -0.50~0.60 | During an endotracheal intubation via direct laryngoscopy, where exposure to aerosolized respiratory particles is at the maximum (e.g., for anesthesiologists) |
| -0.40~0.50 | Exposure to a coughing patient who is not wearing a mask |
| -0.30~0.40 | Doing a nasopharyngeal or oropharyngeal swab on COVID-19 patients |
| -0.20~0.30 | During an endotracheal intubation via video laryngoscopy |
| -0.20~0.30 | Exposure to a coughing patient who is wearing a mask |
| +0.20~0.30 | Wearing surgical masks |
| +0.30~0.40 | Wearing N95 masks |
| +0.20~0.30 | Strict compliance of hand hygiene |
| +0.30~0.40 | Having face and eye shields |
| +0.10~0.20 | Wearing gloves |
| +0.50~0.90 | Wearing a full body biohazard suit |
| ≈1.00 | Absolute protection; either being totally absent from the job or out of shift, or being in full and functional PPE (N95 masks, face and eye shields, gloves, biohazard suit); strict compliance with hand hygiene; having effective engineering controls |

\*this is a sample qualitative point system that can be adjusted or modified depending on the situation.

**Figure 4.**
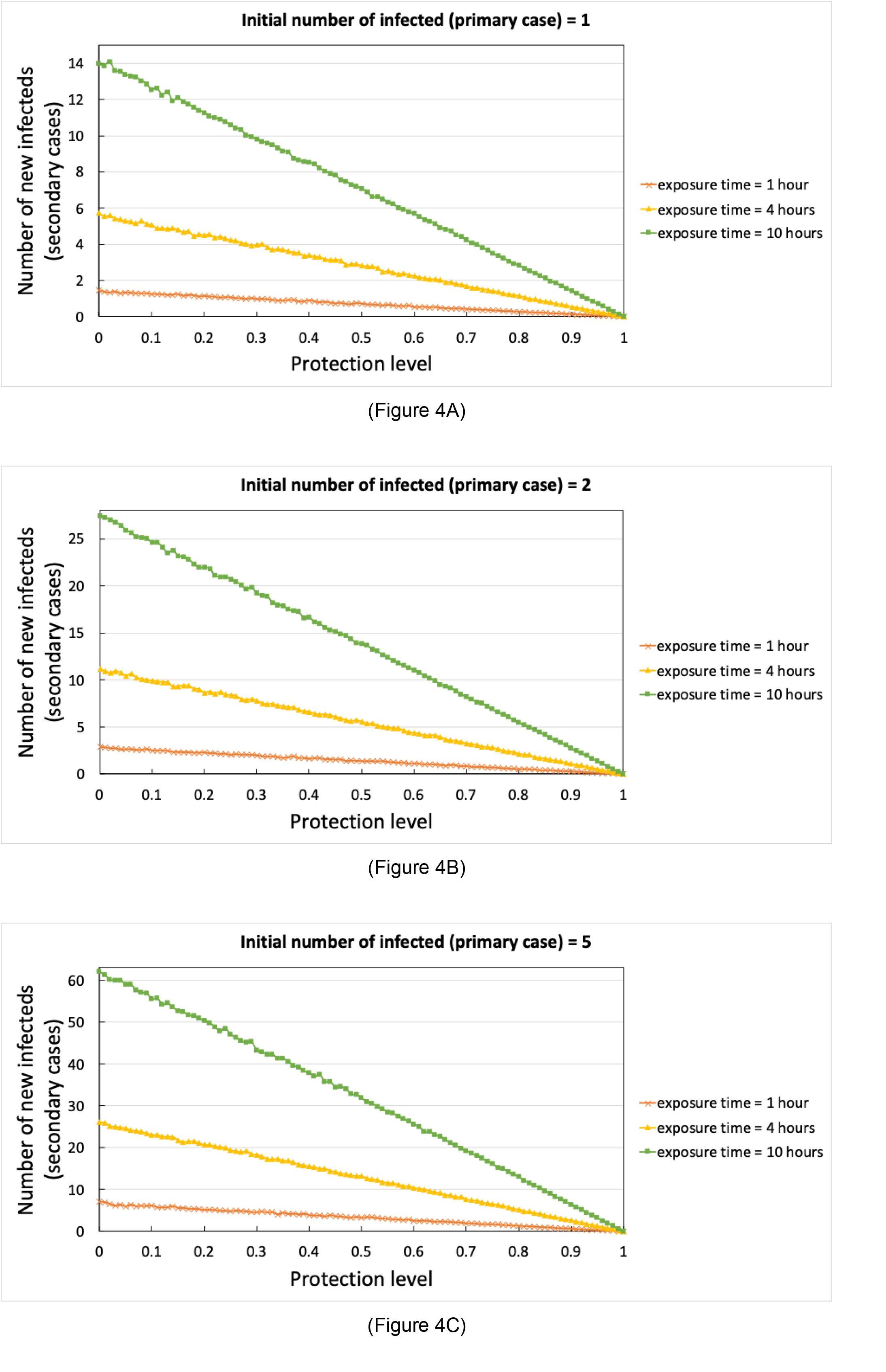
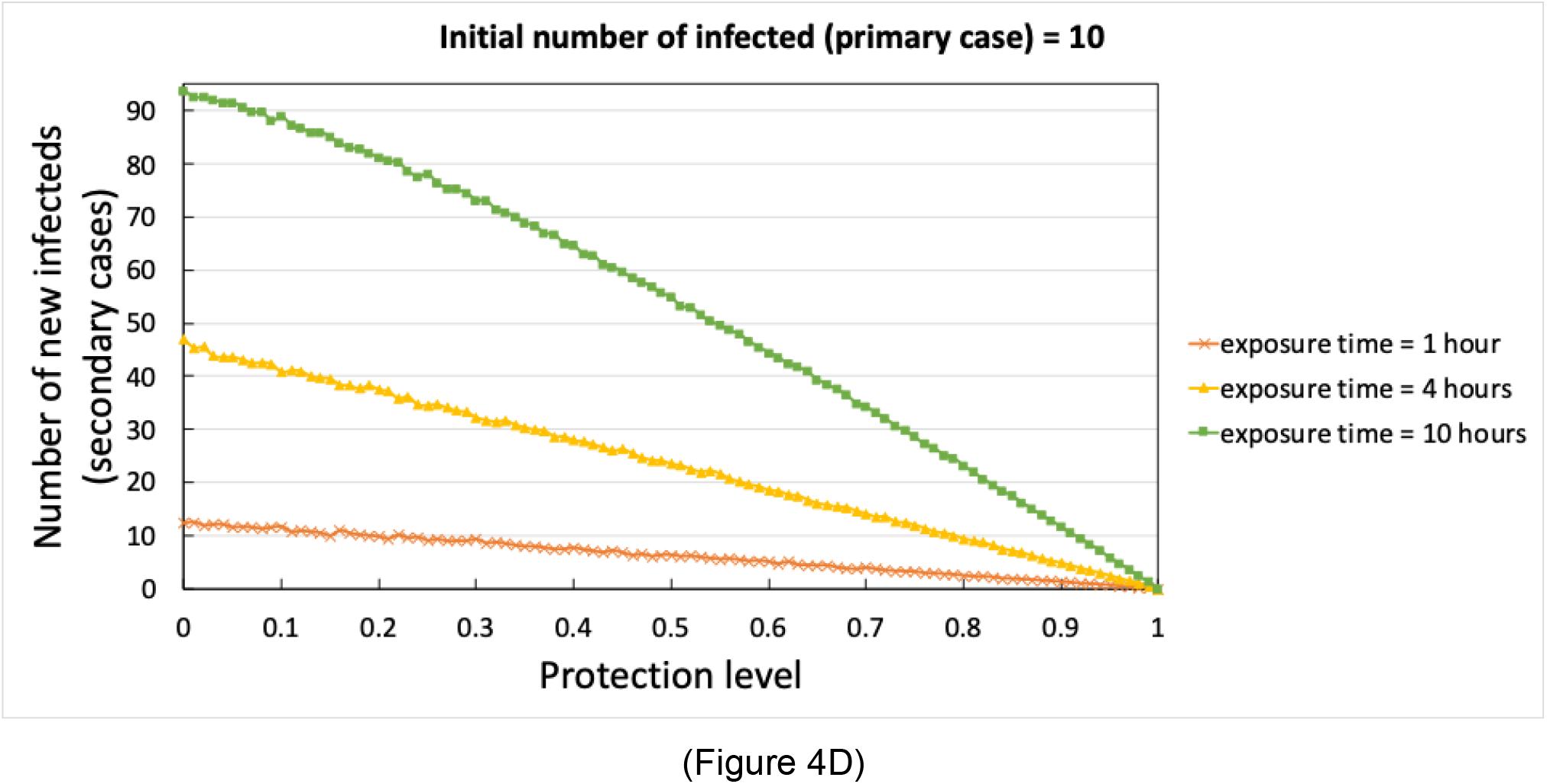
Effect of protection level on the reduction of infection risk. Parameters used: *S*_0_=100, *S*_*max*_=100, no protection, *I*_0_=1, average number of encounters per hour = 120, exposure time = 1 hour. Initial number of infected (**A**) = 1, (**B**) = 2, (**C**) = 5, (**D**) = 10.

Regardless of the number of COVID-19 patients entering a given location at the same time, regardless of the average number of encounters per hour, and regardless of the work shift duration or exposure time, the protection level removes a substantial fraction of the risk faced by the health care worker (Figure 4). In general, having PPEs confers protection towards the health care worker, but certain procedures, especially doing an endotracheal intubation for critically-ill COVID-19 patients, exposes the health care worker to aerosolized particles.

However, the number of COVID-19 patients entering at the same time at a given place influences the level of protection needed (Figure 5). For even an hour of exposure, when ten COVID-19 patients enter at the same time in the same place, the risk of getting infected with 70% level of protection is the same as the risk of getting infected when there is no PPE worn in a room with at most three COVID-19 patients.

**Figure 5.**
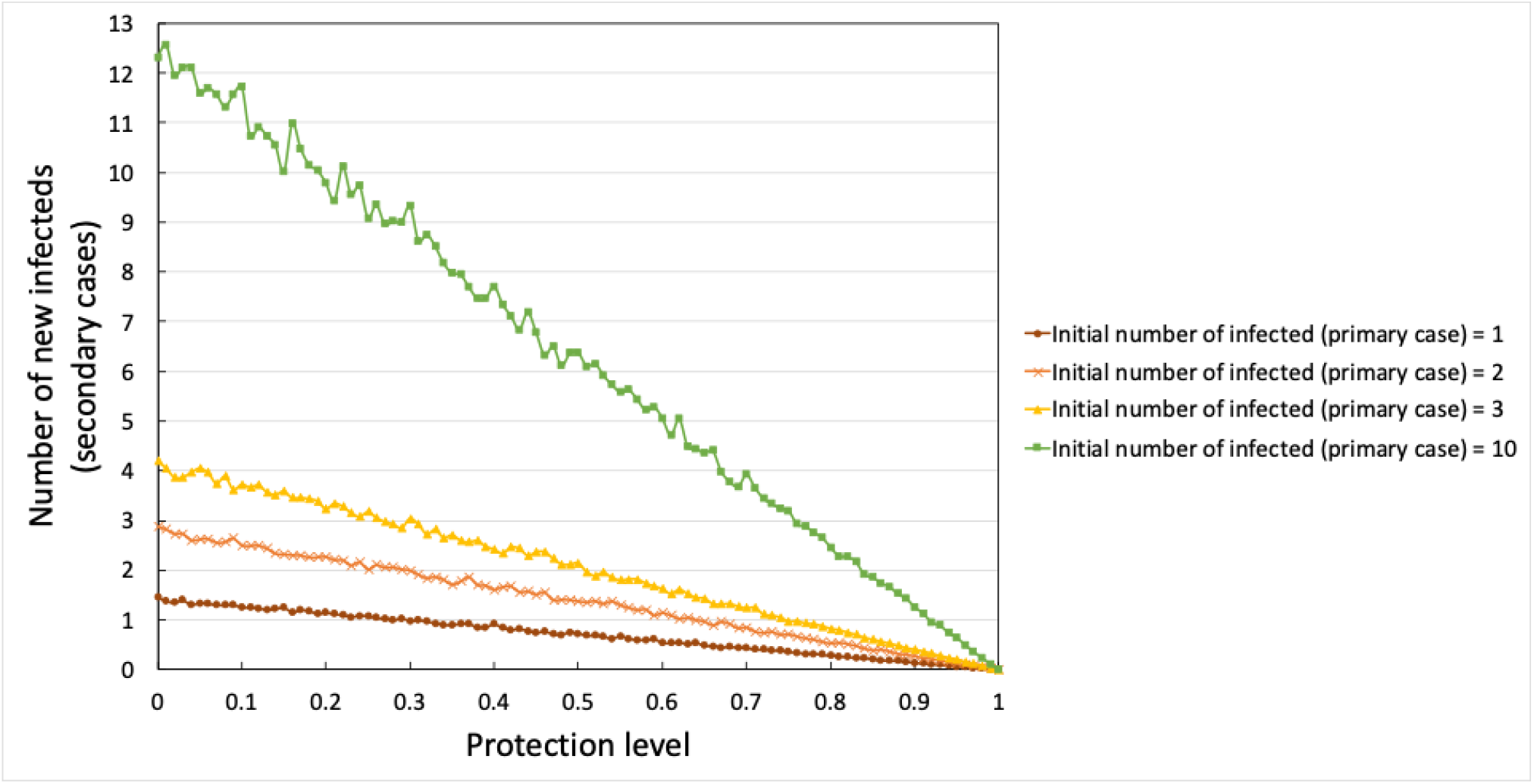
Effect of protection level on reducing the infection risk depending on the number of COVID-19 patients present. Parameters used: *S*_0_=100, *S*_*max*_=100, no protection, *I*_0_=1, average number of encounters per hour = 120, exposure time = 1 hour.

### Risk Assessment

From the previous tables and figures, we therefore propose an overall risk score to be used by each frontliner (Tables 4 and 5). We define the overall risk score as a function of the expected number of new infected persons. If the overall risk score is less than 1.0, then there is a low risk that a person will become infected. An increase in the risk score is proportional to the increase in the number of persons expected to be infected. For example, an overall risk score of 2.00 implies that there are two expected new infected persons. The maximum overall risk score is 10 (where *Points*_*encounter rate*_ =10, *Points*_*duration*_=10, *Crowd Density* =1, and *Protection level*=0). The proposed formula is defined as:

**Table 4.** Proposed risk assessment based on overall risk score

| Overall Risk Score | Risk Assessment |
| --- | --- |
| 0 to less than 0.5 | Low Risk |
| 0.5 to less than 1.0 | Moderate Risk |
| Greater than or equal to 1.0 | High Risk |

**Table 5.** Sample scenarios of frontline health care workers and their corresponding risk score.

| Sample Scenario | Parameter Values | Overall Risk Score (out of 10) |
| --- | --- | --- |
| A PPE-equipped triage nurse in a spacious emergency room of a COVID-19 referral center on an 8-hour shift | $Points_{encounter\ rate} = 10$<br>$Points_{duration} = 2.5$<br>$Crowd\ Density = 0.10$<br>$Protection\ Level = 0.90$ | $\frac{10 + 2.5}{2} \times 0.10 \times (1 - 0.90)$<br>$= 0.0625$<br>**Low Risk** |
| A PPE-equipped anesthesiologist who intubates 1 coughing patient using video laryngoscopy (done in | $Points_{encounter\ rate} = 1$<br>$Points_{duration} = 0.5$<br>$Crowd\ Density = 1.00$ | $\frac{1 + 0.5}{2} \times 1.00 \times (1 - 0.10)$<br>$= 0.675$<br>**Low Risk** |
| a close proximity, lasting for 30 seconds); patient is not wearing PPE | *Protection Level*<br>$= 1.00 - 0.50 - 0.40$<br>$= 0.10$ | |
| A PPE-equipped anesthesiologist who intubates 2 coughing patients using video laryngoscopy (done in a close proximity, lasting for 30 seconds); patient is not wearing PPE | $Points_{\text{encounter rate}} = 2$<br>$Points_{\text{duration}} = 0.5$<br>$Crowd\ Density = 1.00$<br>*Protection Level*<br>$= 1.00 - 0.50 - 0.40$<br>$= 0.10$ | $\frac{2 + 0.5}{2} \times 1.00 \times (1 - 0.10)$<br>$= 1.125$<br>**High Risk**<br>Remark: Note the difference in risk score compared to intubating a coughing patient using video laryngoscopy. |
| A PPE-equipped security guard in a very cramped emergency room of a COVID-19 referral center overwhelmed with patients on a 12-hour shift | $Points_{\text{encounter rate}} = 10$<br>$Points_{\text{duration}} = 6$<br>$Crowd\ Density = 1$<br>*Protection Level* = 0.90 | $\frac{10 + 6}{2} \times 1.00 \times (1 - 0.90)$<br>$= 0.8$<br>**Low Risk (with uncertainty) or Moderate Risk** |
| A PPE-equipped pulmonologist or infectious disease specialist doing quick rounds on 16 patients placed in individual rooms twice during his/her 8-hour shift | $Points_{\text{encounter rate}} = 6.5$<br>$Points_{\text{duration}} = 2.5$<br>$Crowd\ Density = 0.10$<br>*Protection Level* = 0.90 | $\frac{6.5 + 2.5}{2} \times 0.10 \times (1 - 0.90)$<br>$= 0.045$<br>**Low Risk** |
| A security guard wearing only a surgical mask, in a cramped emergency room of a COVID-19 referral center overwhelmed with patients on a 12-hour shift | $Points_{\text{encounter rate}} = 10$<br>$Points_{\text{duration}} = 6$<br>$Crowd\ Density = 1$<br>*Protection Level* = 0.10 | $\frac{10 + 6}{2} \times 1.00 \times (1 - 0.10)$<br>$= 7.2$<br>**High Risk** |
| A patient wearing only a plain surgical mask in a very cramped emergency room together with many other PUIs on queue for 8 hours | $Points_{\text{encounter rate}} = 10$<br>$Points_{\text{duration}} = 5$<br>$Crowd\ Density = 1$<br>*Protection Level* = 0.20 | $\frac{10 + 5}{2} \times 1.00 \times (1 - 0.20)$<br>$= 6.0$<br>**High Risk**<br>Remark: If this patient gets infected, it can compound the risk towards the health workers. |
| A worker wearing a surgical mask in his 6-hour journey back from the Manila to Batangas before the lockdown, riding a cramped jeepney of 10 people, of which 1 is infected, and the infected person is | $Points_{\text{encounter rate}} = 1$<br>$Points_{\text{duration}} = 2$<br>$Crowd\ Density = 1$<br>*Protection Level* = 0.20 | $\frac{1 + 2}{2} \times 1.00 \times (1 - 0.20)$<br>$= 1.2$<br>**High Risk** |
| seated far away from the worker<br>(non-interacting) |  |  |
| A college student, not wearing any PPE, seated beside a COVID-19 infected classmate who is coughing, throughout a 4-hour lecture (interacting) | *Points<sub>encounter rate</sub>* = 10<br>*Points<sub>duration</sub>* = 1.5<br>*Crowd Density* = 1<br>*Protection Level* = 0 | $\frac{10 + 1.5}{2} \times 1.00 \times (1 - 0)$ $= 5.75$ **High Risk** |

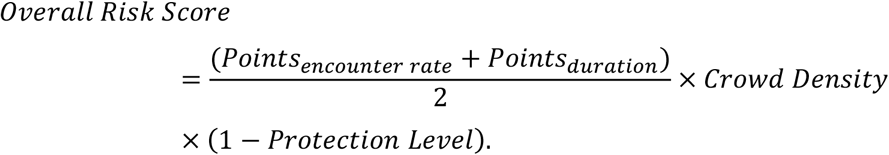

Analogous to the “low” and “high” risk assessments for health care providers of the United States Centers for Disease Control and Prevention (US CDC) [13], we likewise propose a categorization of “low risk” and “high risk”. If the overall risk score is greater than or equal to 1.0, then there is a high risk of a person getting infected.

We can also add an intermediate category between “low” and “high” risks. A “moderate” risk category may be defined as those overall risk scores between 0.5 and 1. By doing a Monte Carlo sensitivity analysis (5 million simulation runs), assuming each input has 10% error, the overall risk score has 0.45 standard error. To account for this uncertainty, overall risk scores between 0.5 and 1 can be classified under the qualitative category of Moderate Risk.

The overall risk score can be used to compare practices. Moreover, as the number of days the health care worker is doing his or her regular job related to a COVID-19 task, the risk of infection increases. The number of days can be scaled accordingly as exposure time.

From the simulations, we have proposed risk assessment criteria and several recommendations. Through these, governments and organizations can have insights on how to minimize or eliminate the transmission of SARS-CoV-2, especially in health care facilities. It is important to protect our health care workers as they are considered essential part of the health care capacity that can provide optimal care to the patients. To test the utility of our model, the model has been used in the Job Risk Profiling Calculator (https://datastudio.google.com/s/iCAEymT7alg) that is used by private and public institutions in the Philippines.

All in all, based on the simulations, the following recommendations can be made:

- Decreasing the rate of patient encounters per frontliner, such as having multiple frontline triage nurses, multiple queues, multiple entrances, and proper referral systems, mitigates the risk of infection. Crowd density factor should always be considered. Having many COVID-19 patients in a room can render a protective measure relatively inadequate. It is recommended to have a quota on the number of COVID-19 patient encounters per duration of work shift. Telemedicine or online consultations can also be an effective control in reducing high risk frontliner-patient interaction [14,15].
- Shorter work shift duration or exposure time reduces the risk faced by the frontliners, especially the security guard and the triage nurse. The protection level against SARS-CoV-2 transmission must be increased accordingly if shortening of work shift duration is not feasible.
- Increased spacing, frequent cleaning of work spaces, and compartmentalizing the rooms of patients in open space decrease the risk of infection not only for health care workers but for the other patients who are COVID-19 negative or non-person-under-investigation (PUI) [16].
- Personal Protective Equipment (PPE) plays a vital role in decreasing the risk of infection, but this protective factor can be overwhelmed by the sheer number of COVID-19 patients (whether confirmed or not). Hence, this should not be relied upon alone, and other structural factors, such as crowd density, be adjusted.
- Frontliners who are handling risky procedures such as endotracheal intubation (i.e., anesthesiologists) on critically-ill COVID-19 patients (who are most likely to be the most infectious), must be given extra protection, and hospital policies must minimize their duration of exposure and the number of patients they encounter or interact with per shift.
- Health care workers play a very important role in a community’s battle against the medical effects of COVID-19. Decreasing the infection risks faced by each health care worker per day, coupled with superior health, well-being and welfare practices, will result in a robust health care staff that can endure a long period of battle during this COVID-19 pandemic.
- Decreasing the infection risk discussed in this paper can also be extended to decreasing the infection risk of non-COVID-19 patients present in a hospital. Moreover, the model and results presented here can be customized for other similar scenarios, such as identifying infection risk in public transportation, school classroom settings, offices, and mass gatherings.
- The recommendations in this paper is based on a theoretical model with parameters calibrated for COVID-19. The theoretical model and algorithm in this paper can be modified for other diseases. It is suggested to validate the results through experiments or cohort and case control studies.

## Conclusion

In this study, we formulated a mathematical model to calculate the risk of being infected in health care facilities. We considered the following factors: (1) the average number of encounters with a suspected COVID-19 patient per hour; (2) interaction time for each encounter; (3) work shift duration or exposure time; (4) crowd density, which may depend on the amount of space available in a given location; and (5) availability and effectiveness of protective gears and facilities provided for the frontline health care workers. A set of risk assessment criteria has been proposed based on the theoretical results to determine if a frontliner is facing low, moderate or high risk of infection.

Based on the simulations and risk assessment, several recommendations are suggested, namely (1) decrease the rate of patient encounter per frontline health care worker, e.g., maximum of three encounters per hour in a 12-hour work shift duration; (2) decrease the interaction time between the frontline health care worker and the patients, e.g., less than 40 minutes for the whole day; (3) increase the clean and safe space for social distancing, e.g., maximum of 10% crowd density, and if possible, implement compartmentalization of patients; and/or (4) provide effective protective gears and facilities, e.g., 95% effective, that the frontline health care workers can use during their shift.

## Data Availability

Codes are included in the manuscript.

## Acknowledgement

JFR is supported by The Abdus Salam International Centre for Theoretical Physics Associate Scheme, Trieste Italy.

## Declarations

The author declares that there are no conflicts of interest.

## Appendix Methods

The estimated values for the number of possible newly infected patients are generated using the Runge-Kutta 4 (RK4) Method of integration to solve the system of differential equations. We use the software (Berkeley Madonna for Mac ver.9.1.19). The differential equations are based on an S-E-I compartment model of disease transmission.

**Figure A1.**
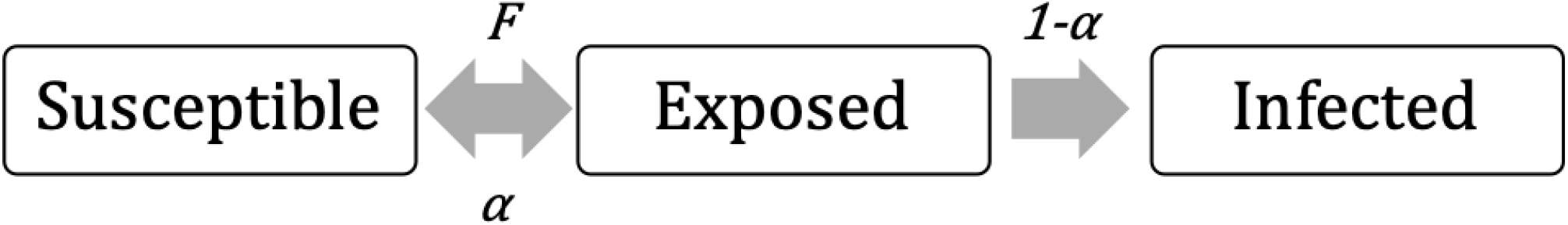
An S-E-I compartment model of disease transmission.

The model is described by the following system of differential equations:

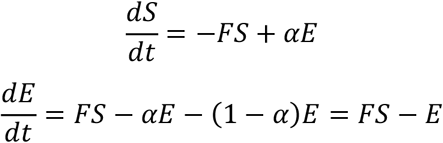

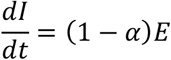

where *S, E, I* are the number of susceptible, exposed, and infected persons (in this case, frontliners); *F* is the force of infection; *α*is the rate of an exposed individual becoming not infected (e.g., through handwashing or other protective measures); and 1−*α*is the rate of getting infected.

The force of infection *F* is defined as:

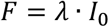

where *λ*is the effective transmission rate, and *I*_0_is the initial number of infected persons (or “the inoculum”), where the effective transmission rate *λ*is defined as:

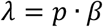

where *β*is the transmission risk or probability, and *p* is the total contact rate. We cannot express *p* as *I/N*, which is the ratio of the total number of infected persons to the total population in a given area because it assumes that everyone is homogenously distributed in a given place. Instead, we note that *p* is in terms of the fraction of the initial number of susceptibles (*S*_0_) over the maximum number of susceptibles that a given area can accommodate (*S*_*max*_), multiplied by the encounter ratio 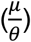, where *μ*is the average number of encounters per hour and *θ*is the threshold number of encounters per hour. Suppose *θ*=*N* per hour. If 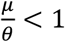 then we are sure that the frontliner has not yet encountered everyone in the room; if 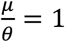 then there is a possibility that the frontliner already encountered everyone in the room; if 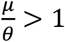 then we are sure that the frontliner encountered a person in the room more than once (by Pigeonhole Principle). The ratio 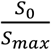 can also be interpreted as the crowd density. The case where *S*_0_=*S*_*max*_and *μ*=*θ*characterizes the usual well-mixed S-E-I model. The parameter *β*(e.g., *β*=0.2[10]) is assumed to be a function of the COVID-19 basic reproductive number (e.g., *R*_0_ = 3) divided by the infectious period (e.g., *τ*=14). We can also interpret 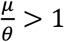 as increasing the average nature of the reproductive number.

Therefore

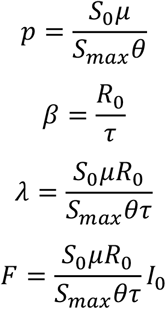

which results in the following:

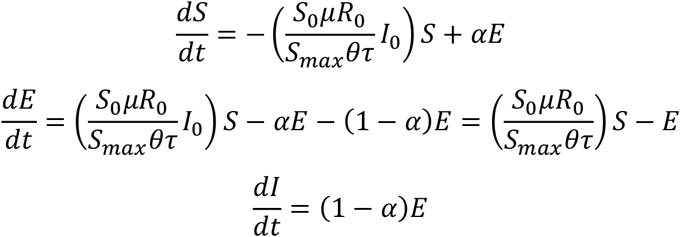

The goal of the study is to investigate the risk of individuals during one cycle of day job. The simulations should end before 48 hours since the dynamics may already change as the newly infected person also becomes infectious.

The exposed (E) class here, in contrast to the classical SEIR model, does not represent persons at the latency stage of the disease. Simply, E class here includes people who have been exposed with the virus (e.g., from respiratory droplets) but the persons can be unexposed through protections (e.g., through washing hands and wearing of face masks). Incubation period is not included because the goal of the study is only for short duration (less than 48 hours). The I compartment is only for counting how many new individuals have been infected, and is not intended to reflect feedback loop leading to a community outbreak. The paper is for risk assessment and not intended to simulate epidemics in the whole community.

The following is the Berkeley Madonna code:

METHOD RK4

STARTTIME = 0

STOPTIME = 2*24;hours

DT = 0.01

;Equations

d/dt (S) = -timer*S*I0*S0*mu*R0/(Smax*N*tau) + alpha*E

crowd_density = S0/Smax

d/dt (E) = timer*S*I0*S0*mu*R0/(Smax*N*tau) - alpha*E - (1-alpha)*E

d/dt (I) = (1-alpha)*E

N = S + E + I0

timer = if TIME>exposure_time then 0 else 1

; you can change TIME>exposure_time to account for the time duration of exposure

R0 = 3+Poisson(superspread)

superspread=1

tau = Normal(14,14*0.1)*24; infectious period times the number of hours in a day limit tau>=0

mu = 1

exposure_time = 1

alpha = 0

init S = S0

S0 = 100

init E = 0

init I = I0

I0 = 1

newinfected = I-I0

limit S<=Smax

limit S>=0

Smax=100

limit E>=0

limit I>=0

; The assumed R0=3 and tau=14 can be changed. The choice of this in the numerical example is based on studies that the force of infection beta=R0/tau is around 0.2 (min of beta=0.05 for asymptomatics, and max of beta=R0/tau=3/6 or 2.5/5=0.5 for symptomatics).

## References

[1] Novel Coronavirus (2019-nCoV) situation reports. (2020). World Health Organization (WHO). https://www.who.int/emergencies/diseases/novel-coronavirus-2019/situation-reports

[2] ztaly coronavirus death toll overtakes China. (2020). BBC. Retrieved 20 March 2020 from https://www.bbc.com/news/world-europe-51964307

[3] Ferguson NM et al. (2020). Impact of non-pharmaceutical interventions (NPIs) to reduce COVID19 mortality and healthcare demand. Imperial College COVID-19 Response Team. https://www.imperial.ac.uk/media/imperial-college/medicine/sph/ide/gida-fellowships/Imperial-College-COVID19-NPI-modelling-16-03-2020.pdf

[4] Anderson RM et al. (2020). How will country-based mitigation measures influence the course of the COVID-19 epidemic? The Lancet, 395: 931–934. https://www.thelancet.com/journals/lancet/article/PIIS0140-6736(20)30567-5/fulltext

[5] Dayrit MM et al. (2018). The Philippines health system review. World Health Organization. Regional Office for South-East Asia. https://apps.who.int/iris/handle/10665/274579

[6] Panela S. (2020). Metro Manila coronavirus cases could peak mid-April if not contained, math model shows. Rappler. Retrieved 22 March 2020 from https://www.rappler.com/nation/255060-math-model-coronavirus-cases-metro-manila-could-peak-mid-april-not-contained

[7] Baticulon R. (2020). OPINION: The Philippine healthcare system was never ready for a pandemic. CNN Philipiines. Retrieved 22 March 2020 from https://cnnphilippines.com/life/culture/2020/3/20/healthcare-pandemic-opinion.html

[8] Rabajante JF. (2020). Insights from early mathematical models of 2019-nCoV Acute Respiratory Disease (COVID-19) Dynamics. Journal of Environmental Science and Management, 23-1: 1–12.

[9] Choudhury A, Kaushik S & Dutt V. (2018). Social-network analysis in healthcare: analysing the effect of weighted influence in physician networks. Network Modeling Analysis in Health Informatics and Bioinformatics, 7: 17.

[10] Ferretti L et al. (2020). Quantifying SARS-CoV-2 transmission suggests epidemic control with digital contact tracing. Science, 368: eabb6936.

[11] Cortez MJV et al. (2017). From epigenetic landscape to phenotypic fitness landscape: Evolutionary effect of pathogens on host traits. Infection, Genetics and Evolution, 51: 245–254.

[12] van Doremalen N et al. (2020). Aerosol and Surface Stability of SARS-CoV-2 as Compared with SARS-CoV-1. New England Journal of Medicine. doi:10.1056/nejmc2004973

[13] United States Centers for Disease Control and Prevention. (2020). Interim Operational Considerations for Public Health Management of Healthcare Workers Exposed to or with Suspected or Confirmed COVID-19: non-U.S. Healthcare Settings. United States Centers for Disease Control and Prevention. https://www.cdc.gov/coronavirus/2019-ncov/hcp/non-us-settings/public-health-management-hcw-exposed.html

[14] Shaikh A. (2015). The impact of SOA on a system design for a telemedicine healthcare system. Network Modeling Analysis in Health Informatics and Bioinformatics, 4: 15.

[15] Burton SH, Tanner KW & Giraud-Carrier CG. (2012). Leveraging social networks for anytime-anyplace health information. Network Modeling Analysis in Health Informatics and Bioinformatics, 1: 173–181.

[16] Nishiura H et al. (2020). Closed environments facilitate secondary transmission of coronavirus disease 2019 (COVID-19). medRxiv. https://www.medrxiv.org/content/10.1101/2020.02.28.20029272v1

